# Methodological Approach for Wastewater Based Epidemiological Studies for SARS-CoV-2

**DOI:** 10.1101/2021.02.17.21251905

**Authors:** Harishankar Kopperi, Athmakuri Tharak, Manupati Hemalatha, Uday Kiran, C. G. Gokulan, Rakesh K Mishra, S Venkata Mohan

**Affiliations:** Bioengineering and Environmental Sciences Lab, Department of Energy and Environmental Engineering (DEEE), CSIR-Indian Institute of Chemical Technology (CSIR-IICT), Hyderabad-500007, India; CSIR-Centre for Cellular and Molecular Biology (CSIR-CCMB), Hyderabad-500007, India; Academy of Scientific and Innovative Research (AcSIR), Ghaziabad, India

**Author notes:** These authors contributed equally to the work (Harishankar Kopperi, Athmakuri Tharak, Manupati Hemalatha, Uday Kiran, C. G. Gokulan).

**Keywords:** Domestic wastewater, Grab sampling, Composite sampling, Sampling station, Frequency

## Abstract

Post COVID-19 outbreak, wastewater-based epidemiology (WBE) studies as surveillance system is becoming an emerging interest due to its functional advantage as tool for early warning signal and to catalyze effective disease management strategies based on the community diagnosis. A comprehensive attempt was made in this study to define a methodological approach for conducting WBE studies in the framework of identifying/selection of surveillance sites, standardizing sampling policy, designing sampling protocols to improve sensitivity, adopting safety protocol, and interpreting the data. The methodology was applied to a community and studied its epidemiological status with reference to occurrence, persistence, and variation of SARS-CoV-2 genome load in wastewater system to understand the prevalence of infection. Hourly and daily grab samples were analyzed and compared with the composite samples over a surveillance window of 7 days. Based on the SARS-CoV-2 RNA copies/L, faeces shedding, and volume of sewage generated the infected individuals and the population who are in active phase in the studied community was estimated.

## 1. Introduction

The persistence and replication of SARS-CoV-2 in the gastrointestinal (GI) tract and shedding through faeces is being established as a transmission route to the environment settings, which eventually discharges to the wastewater/sewage system (Wang et al., 2020 a,b; Xiao et al., 2020b; Zhang et al., 2020; Young et al., 2020; Woelfel et al., 2020; Zahedi et al., 2021; Venkata Mohan et al., 2021). Detection of SARS-CoV-2 genetic material in the sewage/wastewater documented a significant interest among the research fraternity in the framework of wastewater based epidemiological (WBE) studies (Kumar et al., 2020; Venkata Mohan et al., 2021; Hemalatha et al., 2021). Previously, the wastewater functioned as a surveillance system for poliovirus and Aichi virus (Lodder et al., 2012; Asghar et al., 2014). WBE studies also functioned as an important supplement to clinical surveillance in polio eradication and have the potential to inform the epidemiology of COVID-19 (Xagoraraki and O’Brien. 2020; Shaw et al., 2020). Unlike polio, the COVID-19 vaccine reached the mankind, relatively in very short span of time but with some challenges such as their efficacy among different people and time it takes to reach entire population. In this scenario, the testing of the massive population to contain the spread of the virus is a challenge and therefore, an alternative strategy to assess the disease spread and thereby efficiently manage the disease is critical (Xagoraraki and O’Brien. 2020; Shaw et al., 2020; Medema et al., 2020; Venkata Mohan et al., 2021). Wastewater surveillance is an unbiased tool that helps to establish an early-warning system that would be able to monitor the occurrence, spread and, severity of the infection at a community level and therefore help in early preventative measures and allocation of resources to potentially affected areas (Bibby et., 2013 & 2015; Casanova et al., 2015; Brainard et al., 2017; Torrey et al., 2019), which eventually minimize the outbreak and spread (Daughton et al., 2018; Lednicky et al., 2020; Venkata Mohan et al., 2021). Recent reports employed WBE-based approaches to detect SARS-CoV-2 in domestic/sewage wastewater (Wu et al., 2020; Ahmed et al., 2020a; Wurtzer et al., 2020; La Rosa et al., 2020b; Medema et al., 2020; Usman et al., 2020; Hemalatha et al., 2021). Despite the evidence on the persistence of SARS-CoV-2 RNA in wastewater/sewage, the virus transmission to the community from wastewater infrastructure is yet to be established.

The wastewater/sewage complexity, the dilute nature of biomarker in wastewater and, the inability to pinpoint specific locations are some of limitations in establishing quantitative predictions of viral RNA load WBE in (Ahmed et al., 2020b; Mao et al., 2020; Hart et al., 2020). The accuracy of the assessment of viral load among the community is an important pre-requisite of any WBE Studies (Nakamura et al., 2015; Venkata Mohan et al., 2020). Interpretation with a limited number of positive wastewater sample is challenging (Asghar et al., 2014; O’Reilly et al., 2020). Setting minimum standards for surveillance sites, developing a standardized sampling policy, creating laboratory testing protocols to improve sensitivity and minimizing the risk of cross-contamination would be necessary for an informative mode of surveillance (Wu et al., 2020; Xagoraraki and O’Brien., 2020). Several approaches for sampling were used to increase the volume of a sample which might help to identify the virus in wastewater which makes intractable to handle samples and process in laboratories (O’Reilly et al., 2020). The nature, time and frequency of sampling apart from appropriate sampling station will play a decisive role in comprehensively representing the community in order to obtain reliable data. The sampling protocol adopted can also be used to analyze the pattern of viral loads at different time-frequency and finally provide a basis for regular collection at that particular sampling station. Safety understanding in sample collection, handling and processing is needed. Therefore, in this study, a comprehensive attempt was made to establish a methodological approach to represent a community with reference to the prevalence of infection in the framework of WBE studies in terms of identifying/selection of surveillance sites, standardizing sampling policy, designing sampling protocols to improve sensitivity, adopting safety protocol, and interpreting the data.

## 2. Materials and Methods

### 2.1. Surveillance (sampling) station

The selected sampling station receives domestic wastewater from ∼1.8 lakhs people residing in the sub-urban areas including Tarnaka, HMT Nagar, Lalaguda, and Nacharam (and partly Raghavendra Nagar) in Hyderabad, Telangana (State), India. Based on the available population data, the total domestic flow per day was estimated to be 18 million litre per day (MLD). Domestic wastewater samples were collected at the converging (terminal; downstream) point of all the lateral drains that further leads to the Sewage Treatment Plant (STP; 10 MLD capacity; 17.37°N 78.48°E; Nacharam, India) (Fig 1).

**Figure 1:**
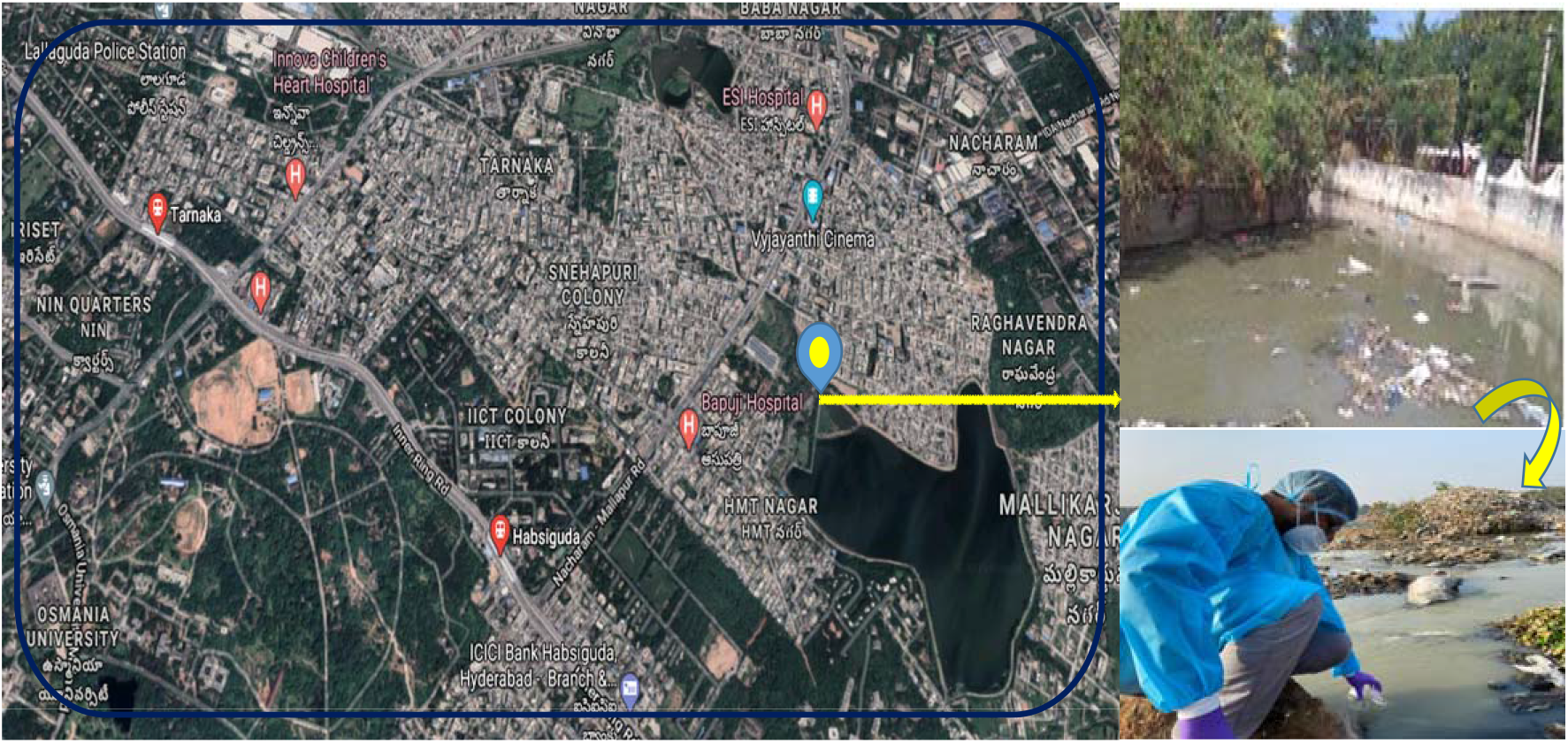
Sampling point selected for daily and hourly monitoring at the inlet of the STP in Nacharam, Hyderabad. (Map source: Google Map).

### 2.2. Sampling Protocol

#### 2.2.1. Type of Samples

The sampling method (grab/composite samples) play a important role on WBE surveillance studies along with population demographics and local epidemiological factors (Alygizakis et al., 2020). An aggregated sample representing an entire community is more to accessible than pooled clinical samples (Murakami et al., 2020). Both grab and composite sampling of domestic wastewater was employed at the selected sampling station in a defined time frequency as detailed in Table 1. A discrete grab sampling was employed at a selected sampling site at multiple time intervals. Viral load, in general, varies over time. Therefore, a composite sample was also prepared by pooling together all grab samples obtained at various hours of the same day or daily samples obtained throughout the sampling week (Table 1). The composite sampling depicts the cumulative viral loads over the selected time-frequency (24 hours or 7 days). The two sampling approaches employed will help mathematically to evaluate the heterogeneity of viral RNA.

**Table 1:**
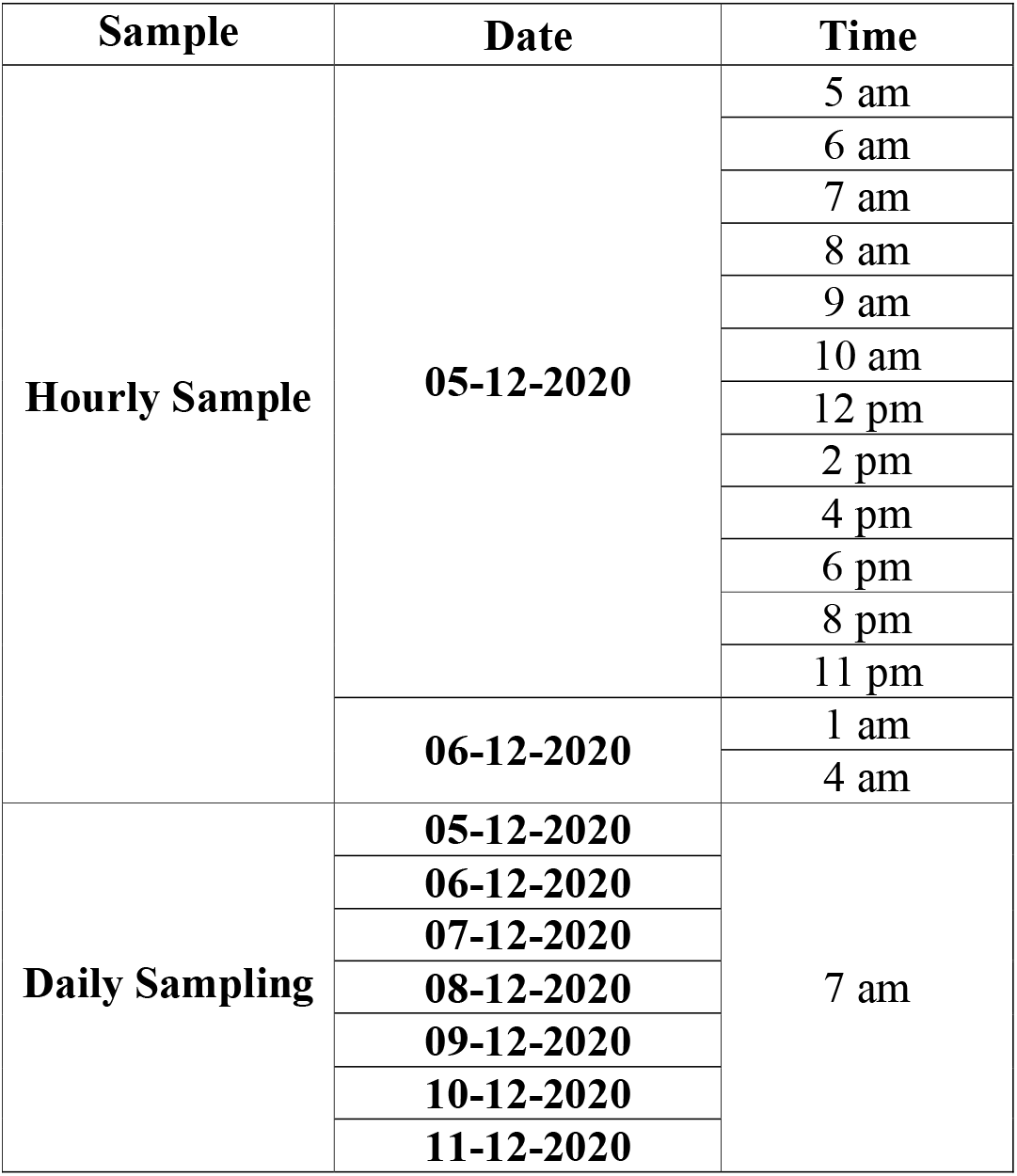
Details of Sampling with reference to time of hourly and daily samples.

**Table 1:** SARS-CoV-2 RNA load with hourly domestic sewage samples.

| Date | Time | E gene* | N gene* | ORF1ab* | RNA copies/L<br>***<br>(using eq. 1) |
| --- | --- | --- | --- | --- | --- |
| 05-12-2020 | 5 am | $28.43 \pm 0.78\%$ | $26.63 \pm 0.40\%$ | $26.98 \pm 3.92\%$ | 22282 |
| | 6 am | $27.18 \pm 3.95\%$ | $25.96 \pm 2.90\%$ | $24.47 \pm 1.14\%$ | 53531 |
| | 7 am | $28.05 \pm 0.39\%$ | $26.88 \pm 2.38\%$ | $26.85 \pm 3.26\%$ | 29086 |
| | 8 am | $28.17 \pm 1.20\%$ | $26.10 \pm 0.34\%$ | $27.07 \pm 0.75\%$ | 26738 |
| | 9 am | $26.31 \pm 5.74\%$ | $25.95 \pm 3.47\%$ | $27.80 \pm 0.85\%$ | 98522 |
| | 10 am | $29.48 \pm 0.98\%$ | $28.34 \pm 1.22\%$ | $28.48 \pm 2.06\%$ | 10671 |
| | 12 noon | $29.17 \pm 0.15\%$ | $27.18 \pm 0.65\%$ | $27.63 \pm 0.10\%$ | 13262 |
| | 2 pm | $29.07 \pm 0.77\%$ | $27.13 \pm 0.55\%$ | $27.24 \pm 0.29\%$ | 14226 |
| | 4 pm | $28.77 \pm 1.08\%$ | $27.55 \pm 1.75\%$ | $27.66 \pm 0.66\%$ | 17556 |
| | 6 pm | $28.79 \pm 0.43\%$ | $27.92 \pm 0.82\%$ | $27.38 \pm 0.60\%$ | 17312 |
| | 8 pm | $29.67 \pm 1.26\%$ | $27.88 \pm 0.29\%$ | $28.02 \pm 1.25\%$ | 9340 |
| | 11 pm | $29.26 \pm 0.27\%$ | $28.07 \pm 0.88\%$ | $27.80 \pm 0.79\%$ | 12451 |
| 06-12-2020 | 1 am | $29.28 \pm 1.09\%$ | $27.62 \pm 0.64\%$ | $27.74 \pm 1.38\%$ | 12278 |
| | 4 am | $30.33 \pm 0.65\%$ | $29.28 \pm 3.09\%$ | $28.88 \pm 1.83\%$ | 5880 |
| **Average** |  | **$28.62 \pm 1.34\%$** | **$27.32 \pm 1.38\%$** | **$27.43 \pm 1.35\%$** | **18292** |

\*Represent $\bar{x} \pm \text{RSD}$ ; \*\*\*RNA copies (based on E gene) were calculated based on the linear fit equation.

#### 2.2.2. Frequency of sampling

A total number of 14 grab samples were collected for hourly monitoring starting from 5 am on 05-12-2020 to 4 am on 06-12-2020 (Table 1). A total of 7 wastewater samples was collected at 7 am from 05-12-2020 to 11-12-2020 for daily monitoring (Table 1). The time period between 5 am to 9 am represent maximum sewage flow at the selected sampling station. No rainfall was recorded during the sampling window of 7 days. The two composite samples were prepared by pooling the hourly and daily collected grab samples individually.

#### 2.2.3. Sampling Procedure & Safety Considerations

The grab samples were collected in a clean plastic container (disposable; 1.2 litres) containing 20 ml of sodium hypochlorite (0.1%) to inactivate the pathogens (Hemalatha et al., 2021). Adequate biosafety measures were taken during sampling and processing of the samples. Samples were collected by wearing PPE kit comprising of gloves, cover suite, eye safety glasses, N95 protective mask and shoes (Hemalatha et al., 2021). For sample collection, the sample container was slightly lowered in the opposite direction of flow with partial immersion. Grab sample volume of one litre was collected at one time with three replicates. Sample information was prepared in the form of field sheets (date and time) and position notified (GPS readings) and point codes, observations. After sampling, the exposed surface of the container was disinfected (isopropyl alcohol (70%)) and sealed in multi-layered plastic covers, labelled, and transported (2-4 °C) to lab and stored at 4°C until further processing. Samples were processed within 12 h after sampling for SARS-CoV-2 detection. Prior to use, all utilities (including PPE kit) are sealed in bio-hazard bag and subjected for sterilization before disposal. The unused samples and materials were inactivated/disinfected before disposal.

### 2.3. Processing of Samples

All the sample processing and detection experiments were performed in a Biosafety level 2 (BSL-2) laboratories as outlined by Hemalatha et al., (2021). Initially, collected samples were filtered using 1 mm filter papers to remove the larger debris followed by secondary filtration using 0.2 µm filtration units (Nalgene® filtration system(vacuum)) to remove other fine particles and pathogens. The filtrate (60 mL) was concentrated to ∼600 µl using 30 kDa Amicon® Ultra-15 (15 ml; Merck Millipore) by ultra-filtration (4 °C; 4000 rpm; 10 min). The concentrated samples (∼600 μL) were aliquoted to 1.5 mL eppendorf vials and 150 μL of the sample was subjected for RNA extraction.

### 2.4. RNA extraction and RT-PCR

The RNA extraction was done using Viral RNA isolation kit (QIAamp, Qiagen) according to the manufacturer’s protocol. Fosun COVID-19 RT-PCR Detection Kit (Shanghai Fosun Long March Medical Science Co., Ltd, China) approved by FDA (Food and Drug Administration, USA) was use to isolate SARS-CoV-2 RNA. The kit consists of primers and probes that target the envelope protein coding gene (E-gene; ROX), nucleocapsid gene (N-gene; JOE), and open reading frame1ab (ORF1ab; FAM) of SARS-CoV-2. The RT-PCR (QuantStudio^TM^5) was performed as per manufacturer recommendations. The reaction program includes reverse transcription (50°C for 15 min) and the initial denaturation (95°C for 3 minutes) followed by 45 cycles at 95°C for 5 seconds and 60°C for 40 seconds. The signals of FAM (ORF1ab), JOE (N gene), ROX (E gene), and CY5 (Internal reference) during the cycling stage was monitored. Positive and negative controls provided along with kit were also included in the reaction plates. All the samples were tested in triplicates. Assessment of RT-PCR kit efficiency, estimation of copy number calculation and virus recovery from sewage were performed as discussed elsewhere (Hemalatha et al., 2021). The filtration units, filter papers and sample bottles were discarded in biosafety bags followed decontamination.

### 2.5. Data Analysis

In order to calculate the number of RNA copies per litre of collected domestic wastewater samples, linear fit equation of the E-gene was employed by which RNA copies per litre wastewater can be calculated (Hemalatha et al., 2021).

Number of RNA copies was calculated based on the C_T_ values of E-gene (Equation 1).

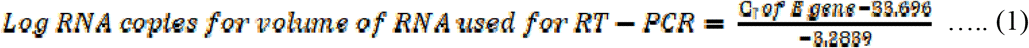

The number of infected people in the given community was calculated based on the average number of RNA copies present in the sewage as per the calculation reported by Ahmed et al., 2020a and Hellmer et al., 2014.

**Method 1 (Ahmed et al**., **2020a)**

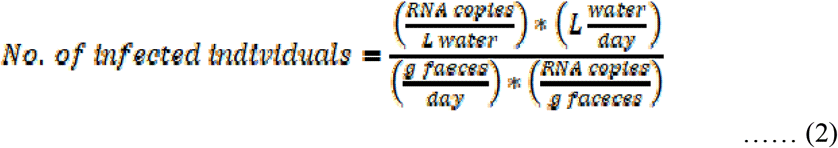

Faeces excreted/person/day = 128 g (Rose et al., 2015). One positive person sheds 10^7^ RNA copies/g of faeces (maximum estimate) (Foladori et al., 2020, Bivins et al, 2020).

**Method 2 (Hellmer et al**., **2014)**

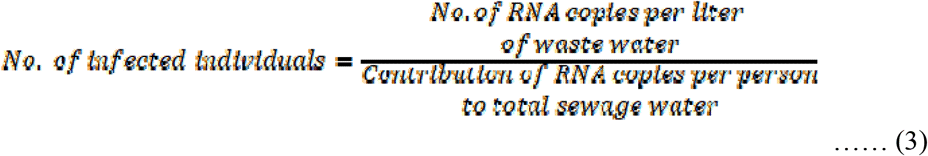

Number of RNA copies excreted per mL of faeces = 10^7^. Volume of faeces excreted = 120 mL (calculated by considering the density of human faeces is 1.07 g/mL) (Foladori et al., 2020).

Relative standard deviation (RSD) for C_T_ value of each gene (E-gene, N-gene, ORF1ab) was calculated using equation 4, where 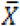 is the mean of C_T_ value and ‘S’ is the standard deviation.

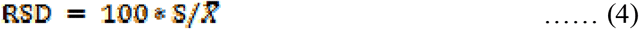

Number of individuals who are in active Phase of Infection during the window period was calculated by considering the infected individuals, window period and infectious period of an infected individual.

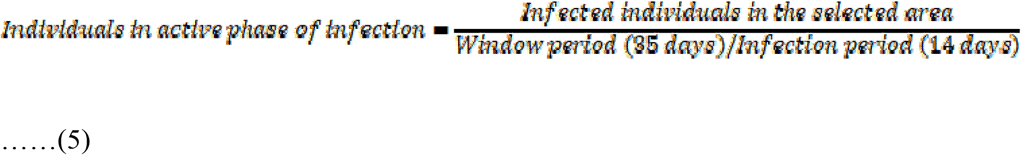

## 3. Results and Discussion

### 3.1. Hourly Sampling for Detection of Viral Load in Sewage

SARS-CoV-2 genetic material was detected in all the hourly samples (n=14) with temporal variation in the viral load. Dynamic detection for viral genome was observed in the domestic sewage indicated the spread of SARS-CoV-2 among the selected community. C_T_ value based on the average of each gene showed 28.62±1.34% for E-Gene, 27.32±1.38% for N-gene and 27.43±1.35% for ORF1ab in the time frame window of 24 hours (Fig.2; Table 1). RNA copy number calculated based on E-gene (Hemalatha et al., 2021) showed a 24-hour average of 22,871 RNA copies/L in the hourly monitored samples (Table 1; Fig 2b). From 5 am to 9 am, the average RNA copies recorded was 45,456 RNA copies /L which was relatively higher compared to the average of 14 hours window (10 am to 1 am; 13,387 RNA copies/L).

**Figure 2:**
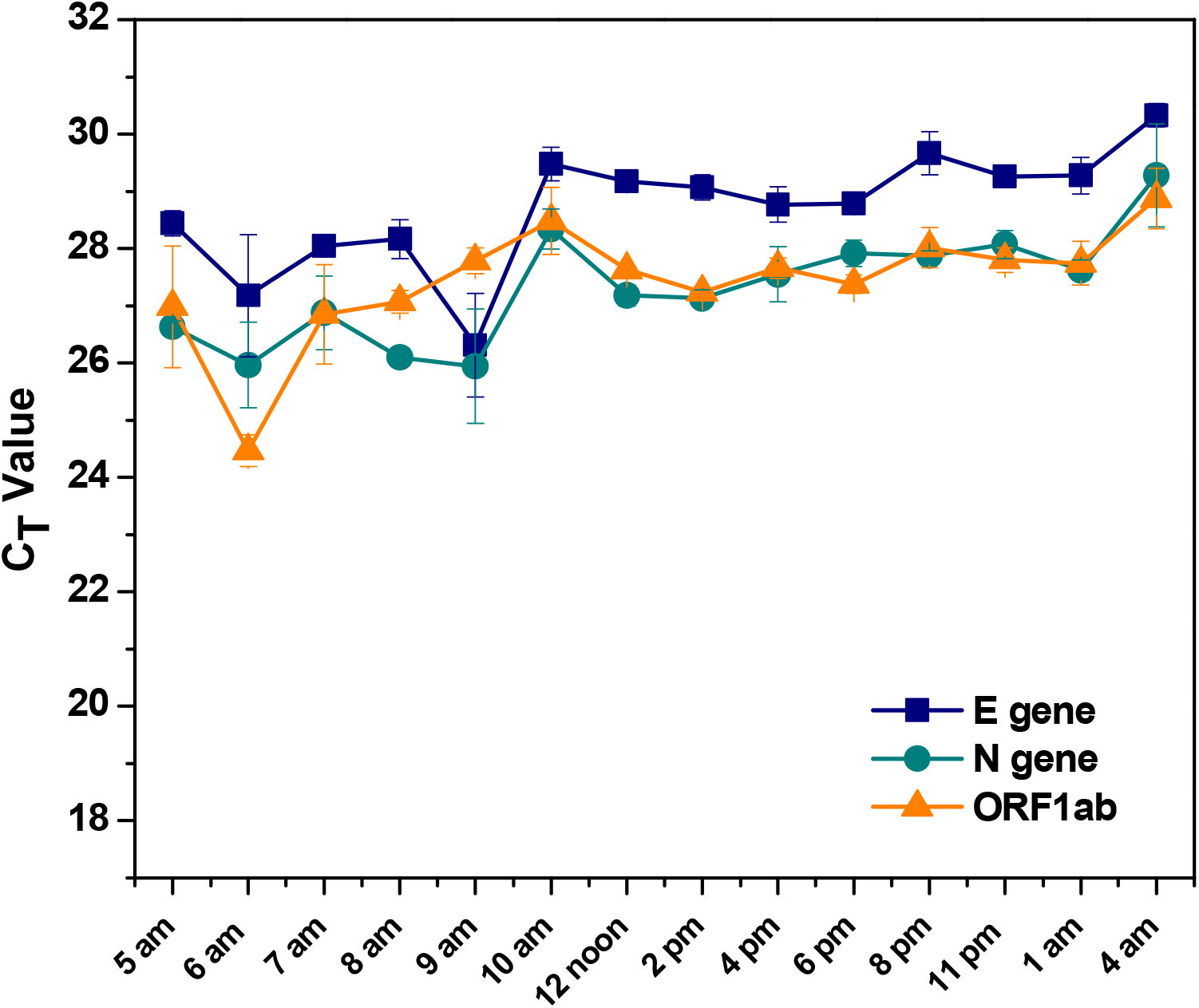
**C**_**T**_ **values of E gene, N gene, and ORF1ab of samples collected hourly from selected point (All values represent**, 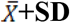).

Higher RNA copies were observed in the samples collected at 6 am (53531 RNA copies/L) and 9 am (98522 RNA copies/L). Between 10 am and 1 am (nearly 14 hours) the viral loads got stabilized between 9,340 and 17,556 RNA copies/L. The lowest viral load of 5,880 RNA copies/L was recorded with the sample collected at 4 am. Individual C_T_ values of three genes also followed the trends with the average C_T_ and RNA copies. E gene C_T_ value varied between 26.31±5.74% and 29.67±1.26%. Similarly, the C_T_ values of N gene and ORF1 ab was observed between 25.95±3.47% to 28.07±0.88% and 24.47±1.14% to 28.48±2.06% respectively. Higher viral load (genetic material) was observed in early hours i.e., 6 am to 9 am, where the peak domestic activity will happen normally during the 24 hours window. Higher detection of viral genome in morning hours might be attributed to the shedding through faeces, which represent the majority of the population in the community. Several considerations like sampling frequency, variation in sampling method, the concentration of disinfectant added, the flow rate of sewage, storage condition, downstream process, etc. effect the RNA copy numbers (Ahmad et al., 2020).

### 3.2. Understanding the dynamics of SARS-CoV-2 RNA load on daily basis

Based on the results obtained from the hourly monitoring, sampling time of 7 am was commonly chosen for daily monitoring to obtain the average distribution of the viral load. SARS-CoV-2 genetic material was detected in all the seven daily monitored samples collected during the window period of one week (05-12-2020 to 11-12-2020) (Table 2; Fig 3). Detection of the viral genetic material over 7 days window period indicated the presence of SARS-CoV-2 RNA in domestic wastewater.

**Table 2:** SARS-CoV-2 RNA C_**T**_ values detected for daily sample.

| Date | Time | E gene* | N gene* | ORF1ab* | RNA<br>copies/L<br>***<br>(using eq. 1) |
| --- | --- | --- | --- | --- | --- |
| 05-12-2020 | 7am | 28.05±0.39% | 26.88±2.38% | 26.85±3.26% | 29086 |
| 06-12-2020 | 7am | 29.20±1.28% | 27.25±0.74% | 27.81±1.13% | 12986 |
| 07-12-2020 | 7am | 31.37±2.80% | 30.35±2.26% | 29.60±0.20% | 2836 |
| 08-12-2020 | 7am | 30.24±1.33% | 29.29±3.58% | 28.67±2.51% | 6263 |
| 09-12-2020 | 7am | 28.72±0.44% | 27.56±0.53% | 27.59±0.30% | 18182 |
| 10-12-2020 | 7am | 27.75±0.26% | 26.03±1.00% | 26.49±0.21% | 35895 |
| 11-12-2020 | 7am | 28.54±0.21% | 26.84±0.73% | 27.19±0.49% | 20628 |
| **Average** |  | **29.12±0.96%** | **27.74±1.60%** | **27.74±1.16%** | **17982** |

\*Represent $\bar{X}$ +RSD; \*\*\*RNA copies (based on E gene) were calculated based on the linear fit equation.

**Figure 3:**
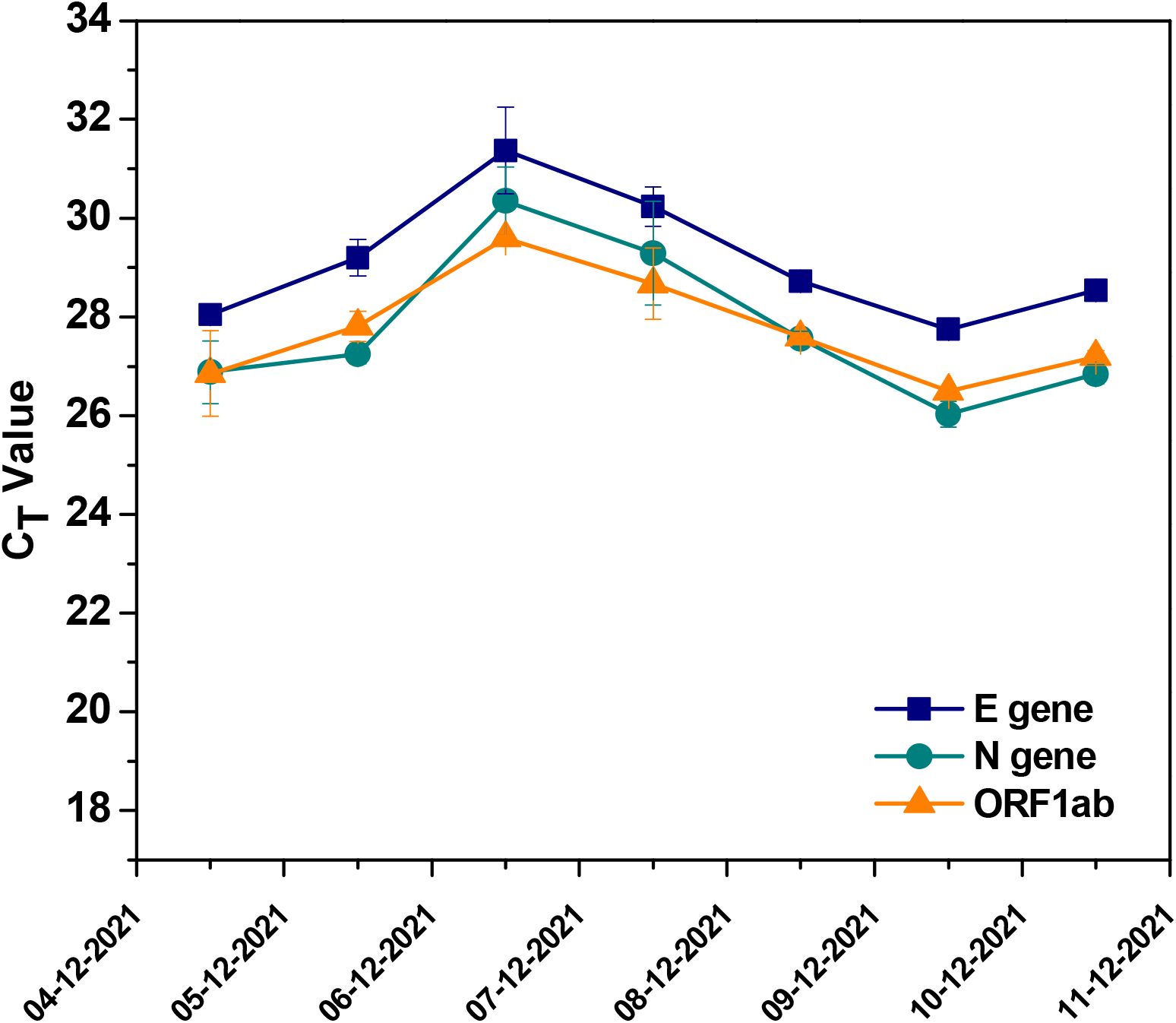
**a) C**_**T**_ **values of E gene, N gene, and ORF1ab of samples collected hourly from selected point (All values represent**, 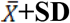).

E-gene C_T_ values varied between 27.75±0.26% and 31.37±2.80%with an average of 29.12±0.96%, N-gene between 26.03±1.00% and 30.35±2.26% with an average of 27.74±1.60% and ORF1ab between 26.49±0.21% and 29.60±0.20% with an average of 27.74±1.16%. Progressive increase in C_T_ values followed by decrement was observed in 7 days of sampling. The RNA copies/L ranged from 2,836 to 35,895 with an average of 17,982.

### 3.3. Composite Sample Analysis

To understand the consistency of sampling procedure two composite samples (hourly and daily) were analysed during the study period (Fig. 4; Table 3). The hourly composite sample was prepared by pooling all the hourly collected samples and similarly for daily composite samples (uniform sample volume). Hourly composite C_T_ of E-gene, N-gene and ORF1ab was detected to be 28.80±1.10%, 27.04±1.43% and 27.56±3.20% respectively. which is correlating well with the C_T_ values of the hourly sample (Table 2; Fig 4). Daily composite C_T_ of E-gene, N-gene and ORF1ab was detected to be 28.62±0.60%, 27.25±1.38% and 26.99±2.58% respectively. which is correlating well with the C_T_ values of the daily sample (Table 2; Fig 4). Quantitatively, RNA copies present in the hourly and daily composite samples was observed to be 19,503 RNA copies/L and 17,191 RNA copies/L, respectively, which correlates well with the average RNA copies of hourly and daily monitored samples 22,871 RNA copies/L and 17,982 RNA copies/L, respectively. Correlation between the composite and cumulative RNA copies represents the efficiency of sampling frequency.

**Table 3:** SARS-CoV-2 RNA load with hourly and daily composite domestic sewage samples.

| Sample | E gene* | N gene* | ORF1ab* | RNA copies/L<br>***<br>(using Eq. 1) |
| --- | --- | --- | --- | --- |
| Hourly Average | 28.71±1.25% | 27.32±1.79% | 27.43±1.35% | 22871 |
| Hourly Composite | 28.80±1.10% | 27.04±1.43% | 27.56±3.20% | 19503 |
| Daily average | 29.12±1.00% | 27.74±1.62% | 27.74±1.15% | 17982 |
| Daily Composite | 28.62±0.60% | 27.25±1.38% | 26.99±2.58% | 17191 |
| **Average** | **28.81±0.99%** | **27.34±1.56%** | **27.43±2.07%** | **19387** |

\*Represent $\bar{x} \pm RSD$ ; \*\*\*RNA copies (based on E gene) were calculated based on the linear fit equation.

**Table 4:**
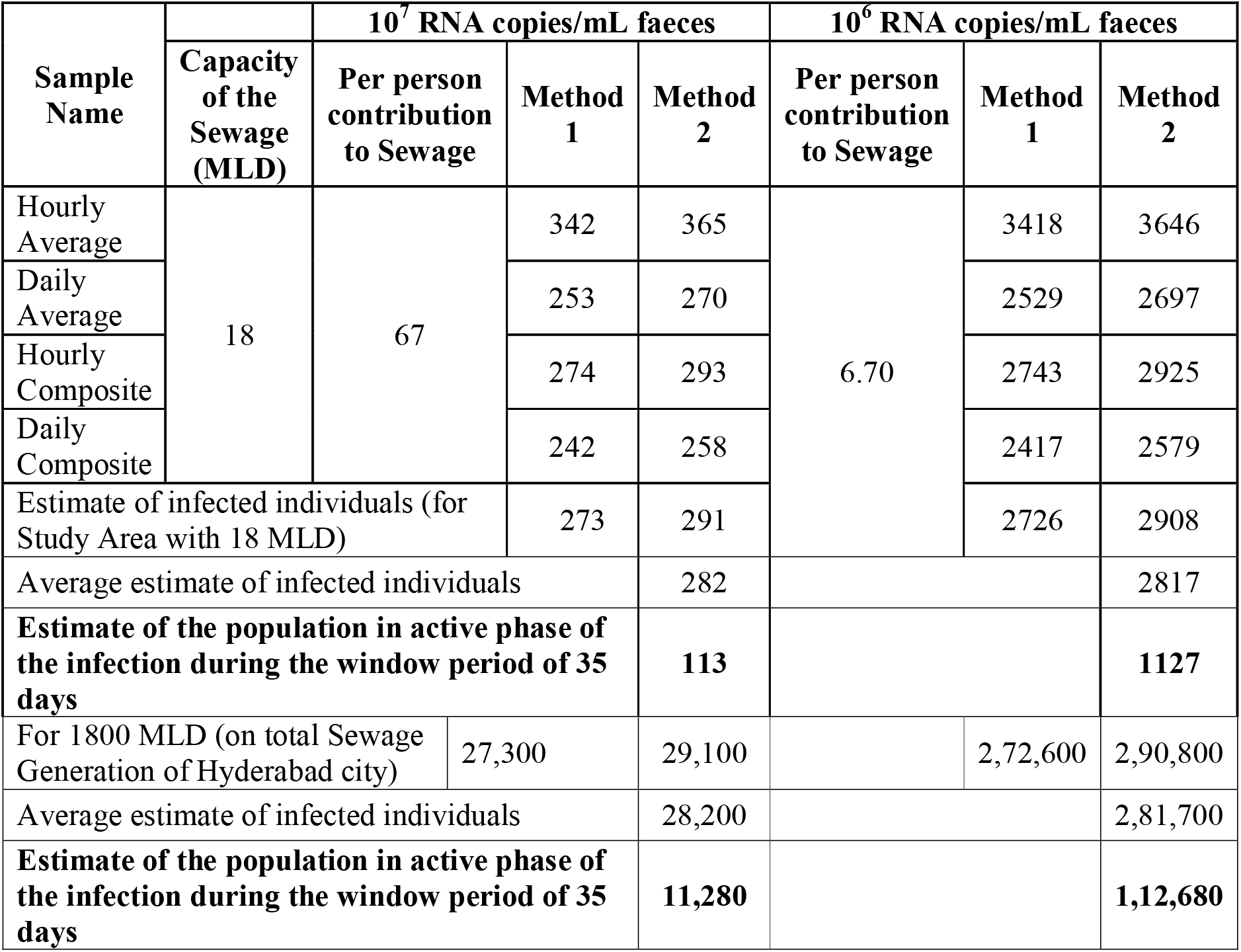
Infection estimating through the number of people infected (symptomatic, asymptomatic, and recovered) during the sampling window on average basis of sampling.

**Figure 4:**
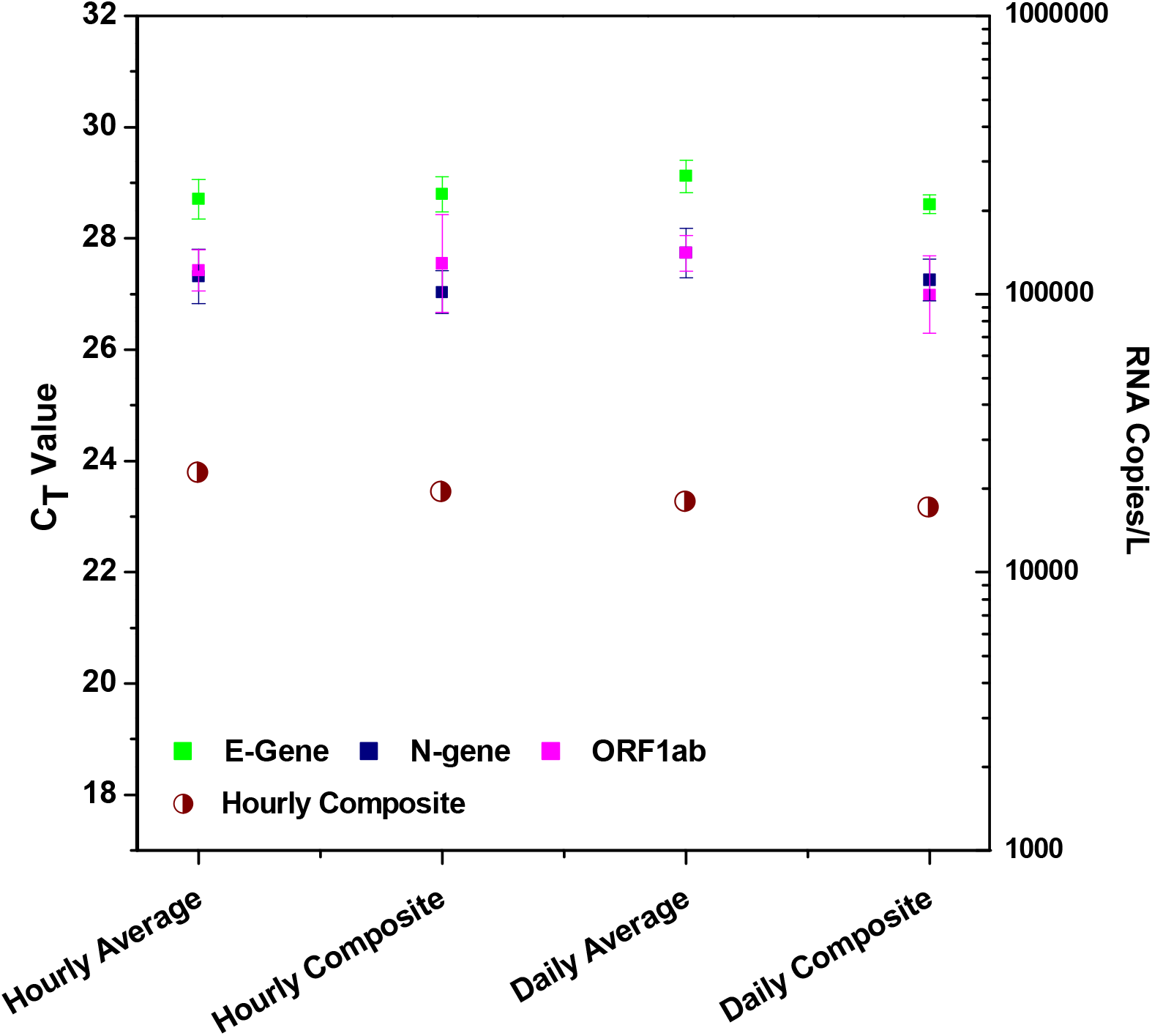
**a) C**_**T**_ **values of E gene, N gene, and ORF1ab and RNA copies of hourly, daily average and composite samples (All values represent**, 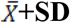).

### 3.4. Epidemiological status- Estimation of Community Infection

To estimate the community spread of the virus, the average RNA copies of hourly average, daily average, and composites (hourly and daily) representing 7 days window of sampling period was taken into consideration along with the capacity of sewage generated in the selected community (Table 5). Both hourly and daily samples showed variability in the viral load with the time scale. The estimated number of infected individuals would include those in the early as well as later stages of infection and shedding viral particles in their faecal matter. This period was chosen based on the reports on persistence of SARS-CoV-2 RNA material for up to 47 days after the infection (Wu et al., 2020). Independent reports highlighted the replication of SARS-CoV-2 in GI tract and the prolonged shedding viral material through faeces during and after active infectious phases (Ahmed et al., 2020a; Kitajima et al., 2020; Holshue et al., 2020; Wu et al., 2020; Wurtzer et al., 2020; Cai et al., 2020; Ling et al., 2020; Woelfel et al., 2020; La Rosa et al., 2020; Xiao et al., 2020a,b). The number of infected individuals were estimated based on shedding range of 10^6^ and 10^7^ RNA copies/mL faeces (Foladori et al., 2020) employing the average RNA copies obtained from this study to avoid the ambiguity/discrepancy in the number of viral particles excreted by infected individuals. With reference to the studied community, the estimated infected individuals during the sampled window period is between 282 (10^7^ RNA copies/mL faeces) and 2817 (10^6^ RNA copies/mL faeces) and the number of individuals in the active phase of infection might be between 113 to 1127 with a wastewater flow rate of 18 MLD. Based on this number the infected individuals for the metropolitan Hyderabad city were also calculated with total sewage generation of 1800 MLD (https://numerical.co.in/numerons/collection/5e8fd4f6f3c42b5803c09a3b).

The number of infected individuals during the sampling window was estimated between 28,200 and 2,81,700 with a active phase of infection between 11,280 and 1,12,680. The loss of 0.02 to 3000 RNA copies/mL was reported during the transit of faeces from point of excretion to the drain (Foladori et al., 2020) which could influence the overall estimation of infected individuals. The obtained figures might include pre-symptomatic, post-symptomatic, asymptomatic, and symptomatic patients of which asymptomatic individuals might be the major contributors based on the community behaviour and serological data. WBE can quantify the scale of infection prevailing in the selected community with a benefit of detection for the individuals who have not been tested, asymptomatic, potentially symptomatic, pre-symptomatic, or only have mild symptoms (Osuolale et al., 2017; Medema et al., 2020; Lodder and de Roda Husman, 2020; Hata and Honda, 2020; Mallapaty, 2020; Naddeo and Liu, 2020; Qu et al., 2020; Venkata Mohan et al., 2021; Hemalatha et al., 2021). Asymptomatic and symptomatic cases also result in significant uncertainty in the estimated extent of SARS-CoV-2 infection (Li et al., 2020). However, this kind of estimation using the C_T_ value and RNA copies could help to predict the near precise number of infected individuals in a selected community/area.

Based on the outcome from this study, the following points can be considered for planning WBE (a) Sample/sampling station(s) should be a true representative of the selected community to be studied; (b) Sampling station(s) should be selected at the downstream converging point of discharge line if flowing water is being analysed. Inlet discharge point to STP can be considered for sampling which comprehensively indicates the infection of the community/area under the STP coverage; (c) Combined grab and composite sampling strategies can be adopted with a define sampling frequency; (d) Sampling frequency should be extended for not less than 24 h for hourly sampling and for not less than 7 days for daily sampling to get a true average representation of the community. For daily sampling, the time of sample should be carefully chosen to represent average values and (e) Estimation of the viral infection of the study area can be calculated based on the population and its wastewater discharge data.

Given the wide-spread transmission of SARS-CoV-2 it is almost impossible to test every individual. Sewage based surveillance is a holistic and effective approach to study the infection dynamics, which helps in the efficient management of the SARS-CoV-2 spread. A clear connection between the RT-PCR quantitative data and the recorded clinical data can be correlated to reduce ambiguity viral-wastewater surveillance (O’Reilly et al., 2020; Xiao et al., 2020a).

Longitudinal sampling from the same location along with metagenomics can provide an additional illustration on the status (Alygizakis et al., 2020). Associated with clinical data, WBE could able to provide information on SARS-CoV-2 transmission within a community including the beginning, tapering, or reemergence of an epidemic (Ahmed et al., 2020b). During the current pandemic, various mutation of SARS-CoV-2 may be evolved and WBE can also facilitate to detect the mutants by employing metagenomic analysis.

## 4. Conclusions

Overall, this study provides a methodological framework for WBE studies towards viral surveillance in wastewater/sewage infrastructure to precisely represent a selected community with a defined window period in terms of identifying/selection of surveillance sites, standardizing sampling policy, designing sampling protocols to improve sensitivity, adopting safety protocol, and interpreting the data. Wastewater/sewage-based surveillance can help to understand the occurrence and spread of the pandemic in the selected population or area in a more compressive approach by adopting a well-defined sampling protocol. Past experience with viral outbreaks there is more chances of epidemic/pandemic outbreaks in future due to the persistent anthropogenic induce ecological disturbances. In the environmental monitoring system, induction of viral/pathogen surveillance as a regular parameter along with routine monitoring of wastewater quality parameter will form an important basis to understand the early warning of the outbreaks. It is strongly recommended that healthcare/environmental agencies include WBE studies periodically to fight present and future pandemic outbreaks. Apart from an early warning signal for predicting the outbreaks, WBE also supports clinical scrutiny along with the disease detection and management systems. The current study also offers a methodological approach to monitor other pathogens. A policy in the framework of public health by appending to the environmental systems will eventually help to safeguard the community with future outbreak.

## Data Availability

The data related to the pre-print is included in the submitted file

## Declaration of competing interest

The authors declare that they have no known competing financial interests or personal relationships that could have appeared to influence the work reported in this paper.

## Acknowledgments

The work was supported by Council of Scientific and Industrial Research (CSIR), New Delhi, India in the form of project entitled ‘Testing for COVID-19 in wastewater as a community surveillance measure (6/1/COVID-19/2020/IMD)’.s UK thanks UGC, CGG and MH thank CSIR for the financial support received. KH, AT, MH and SVM acknowledge the Director, CSIR-IICT for the support.

## Notes

### Competing Interest Statement

The authors have declared no competing interest.

### Author Declarations

Acquired from CSIR-IICT

